# Manual ability in hand surgery patients: validation of the ABILHAND scale in four diagnostic groups

**DOI:** 10.1101/2020.07.02.20144147

**Authors:** Ghady El Khoury, Olivier Barbier, Xavier Libouton, Jean-Louis Thonnard, Philippe Lefèvre, Massimo Penta

## Abstract

**Background:** Patients treated in hand surgery (HS) belong to different demographic groups and have varying impairments related to different pathologies. HS outcomes are measured to assess treatment results, complication risks and intervention reliability. A one-dimensional and linear measure would allow for unbiased comparisons of manual ability between patients and different treatment effects.

**Objective:** To adapt the ABILHAND questionnaire through Rasch analysis for specific use in HS patients and to examine its validity.

**Methods:** A preliminary 90-item questionnaire was presented to 216 patients representing the diagnoses most frequently encountered in HS, including distal radius fracture (n=74), basal thumb arthritis (n=66), carpal tunnel syndrome (n=53), and heavy wrist surgery (n=23). Patients were assessed during the early recovery and in the late follow-up period (0-3 months, 3-6 months and >6 months), leading to a total of 305 assessments. They rated their perceived difficulty with queried activities as impossible, difficult, or easy. Responses were analyzed using the RUMM2030 software. Items were refined based on item-patient targeting, fit statistics, differential item functioning, local independence and item redundancy. Patients also completed the QuickDASH, 12-item Short Form Survey (SF-12) and a numerical pain scale.

**Results:** The rating scale Rasch model was used to select 23 mostly bimanual items on a 3-level scale, which constitute a unidimensional, linear measure of manual ability with good reliability across all included diagnostic groups (Person-Separation Index = 0.90). The resulting scale was found to be invariant across demographic and clinical subgroups and over time. ABILHAND-HS patient measures correlated significantly (p<0.001) with the QuickDASH (r=-0.77), SF-12 Physical Component Summary (r=0.56), SF-12 Mental Component Summary (r=0.31), and pain scale (r=-0.49).

**Conclusion:** ABILHAND-HS is a robust person-centered measure of manual ability in HS patients.

## Introduction

In hand surgery (HS), as in other medical specialties, outcome evaluations are needed to assess the effectiveness and reliability of the intervention, as well as to reinforce patient education regarding the risks and outcomes of the procedure and, potentially, to justify therapeutic practices to payers [1]. Physician-documented reports of HS outcomes based on clinical examination and imaging should be complemented with patient reported outcomes assessed by questionnaires designed to capture patients’ perspectives with respect to the impact of their conditions and interventions on their daily lives [2–4].

Current views of health and disability have been shaped by the World Health Organization’s International Classification of Functioning, Disability, and Health [5], which parses disease consequences into three domains: impairment of anatomical structures (e.g. bones, muscles, ligaments) or body functions (e.g. motor skills, sensitivity), activity limitations (e.g. manual activities), and participation restrictions (e.g. in hobbies and work). The impact of a pathology or a surgical intervention in these three domains is also conditioned by personal factors (motivation, capacity to develop compensatory strategies) and environmental factors (social or professional context). Although impairment measurements such as imaging can provide clues regarding functional prognosis, it does not provide good information about performance in everyday life, especially of the hands, which are important for a great variety of activities [6–8]. For example, demonstration of a bone fracture union is insufficient to determine whether a patient is capable of resuming their usual activities or returning to work [9–12].

The patient-reported questionnaires that have been most commonly used in HS [13] are the Disability of the Arm, Shoulder and Hand questionnaire (DASH) [14], the Patient Rated Wrist Evaluation (PRWE) [15], and the Carpal Tunnel Questionnaire (CTQ) [16]. Each of these questionnaires has been reported to have good psychometric properties, but each has a particular focus on its own area(s) of disablement. The DASH assesses body functions, activities, and participation [17] and can be divided into 3 subscales based on dimensionality [18]. Meanwhile, the PRWE is specific to the wrist joint and the CTQ is specific to carpal tunnel syndrome (CTS). The Michigan Hand Outcomes Questionnaire (MHQ) [19] is a multidimensional hand-specific outcomes instrument consisting of six subscales, measuring overall hand function, activities of daily living, pain, work performance, aesthetics, and satisfaction. It measures impairment by hand (left and right separately), rather than overall disability. Interpretation of total scores on multi-dimensional instruments can be less than straightforward given that patients can show simultaneous improvement in one domain with deterioration in another [20,21]. Assessment of functional recovery on a unidimensional [22] and linear [21] scale would allow for quantitative comparisons of ability among different patients and treatments. Such a scale can be developed with state-of-the-art psychometric methods, such as the Rasch model [23,24].

The ABILHAND questionnaire is a Rasch-model built measure of manual ability [25] that provides an invariant linear scale and allows for quantitative comparisons of manual ability between patients and over time. The scale has been validated in populations with rheumatoid arthritis [26], chronic stroke [8], pediatric cerebral palsy [27], systemic sclerosis [28] and neuromuscular diseases [29]. These previous validations have shown that the difficulty of most manual activities was diagnosis-dependent [30]. Therefore, the objective of this work was to adapt the ABILHAND scale to the most frequent diagnoses treated in HS.

## Methods

### Questionnaire adaptation to HS patients

The ABILHAND is a measure of manual ability that assesses one’s ability to manage daily activities requiring upper limb use, regardless of strategy [25]. The necessary permissions were obtained from the developer of the original questionnaire to modify it. To develop a HS-adapted ABILHAND, a preliminary item list was compiled from previous versions of the ABILHAND questionnaire, the DASH, PRWE, CTQ, and MHQ items together with some new items. This pool of items was submitted to nine HS experts (hand surgeons, physical medicine and rehabilitation physicians, physical therapists, and occupational therapists), who were asked to assess each item’s relevance to hand surgery patients on a yes/no basis and propose additional items that might be affected by the relevant pathologies (e.g. sensation for CTS and wrist loading for distal radius fractures (DRF)). A final list of 90 items constituted the experimental ABILHAND-HS questionnaire (S4 Appendix).

### Patients

A convenience sample of 216 patients was recruited from February 2018 to February 2019 at the HS consultation center at Cliniques Universitaires Saint-Luc, Belgium representing the following four diagnostic categories: CTS, DRF, basal thumb arthritis (BTA), and heavy wrist surgery (HWS, including 1^st^ row carpectomy and partial or total wrist arthrodesis). The inclusion criteria for patients were being >18 years old and being able to read and understand French. The exclusion criteria included comorbidities that may impede manual ability substantially (i.e. tremor, paralysis and active rheumatologic disease) and any mental or cognitive dysfunction (i.e. dementia and mental retardation). The patient characteristics are summarized in Table 1. Patients provided written informed consent to participate. This study was approved by the ethical committee of Cliniques Universitaires Saint-Luc-Université catholique de Louvain (N° B403201523492).

**Table 1.** Sample characteristics (n = 216).

| Characteristic | N (%)<sup>a</sup> |
| --- | --- |
| **Gender** |  |
| Women | 145 (67%) |
| Men | 71 (33%) |
| **Mean age (range), years** | 60.3 (19–93) |
| **Education level** |  |
| Basic | 109 (51%) |
| Postsecondary | 107 (49%) |
| **Work status** |  |
| Student | 2 (1%) |
| Unemployed | 22 (10%) |
| (Self-)Employed | 83 (38%) |
| Retired | 109 (51%) |
| **Hand dominance** |  |
| Right | 194 (90%) |
| Left | 15 (7%) |
| Ambidextrous | 7 (3%) |
| **Involved dominant hand** |  |
| Yes | 136 (63%) |
| No | 80 (37%) |
| **Diagnostic group** |  |
| Distal radius fracture (DRF) | 74 (34%) |
| Basal thumb arthritis (BTA) | 66 (31%) |
| Carpal tunnel syndrome (CTS) | 53 (24%) |
| Heavy wrist surgery (HWS) | 23 (11%) |
| **Follow-up assessments (n = 305)** |  |
| 0–3 months<br>(57 DRF, 52 CTS, 22 BTA, 1 HWS) | 132 (43%) |
| 3–6 months<br>(38 DRF, 16 CTS, 3 BTA, 1 HWS) | 58 (19%) |
| >6 months<br>(30 DRF, 18 CTS, 46 BTA, 21 HWS) | 115 (38%) |

<sup>a</sup>Except where otherwise indicated

### Procedures

The French-language experimental ABILHAND-HS items were presented in five random orders to avoid a systematic item sequence bias. Patients were asked to indicate their perceived difficulty associated with completing the activities without technical or human assistance, independent of the hand used to perform the activity on a three-level scale: impossible (0), difficult (1), or easy (2) [8]. Activities not attempted during the last week were treated as missing responses. Patients also completed the QuickDASH [31], 12-item Short Form Survey (SF-12) questionnaire [32] and a 10-level numerical pain scale, for external validation purposes.

Patients were first assessed as soon as they presented to their hand surgery consultation appointments and had experienced manual activities in their own environment: after hand surgery and cast removal for DRF, BTA and HWS and at the first consultation for non-operated CTS and BTA. For the first assessment, patients were interviewed by the principal investigator in order to ensure clarity, obtain feedback from participants, and make sure instructions are properly followed. Patients were also asked to suggest additional items they felt the questionnaire was missing. However, these were either gender related (e.g. fastening a bra) or very specific and were thus not retained. Follow-up assessments were completed in our consulting office or returned by mail, leading to a total of 305 completed assessments, which provides sufficient power to support the planned Rasch analysis [33].

### Rasch analysis

The 90-item experimental ABILHAND-HS questionnaire responses were analyzed using the Rasch model in RUMM2030 software (RUMM Laboratory Pty Ltd., Perth, Australia). The Rasch model [24], a prescriptive model, requires that specified response probabilities depend on only item difficulty and patient ability. Polytomous datasets with thresholds between successive response categories can be analyzed with either a rating scale model that constrains all threshold locations to be equal across items [34] or a partial credit model that allows threshold locations to vary across items [35]. Patient abilities and item difficulties are located along a common linear, unidimensional continuum that defines the latent variable of interest (i.e. manual ability). The locations are expressed in logits, calculated as the logarithm of the pass/fail probability ratio of an item or threshold. The logit locations were converted into centiles to facilitate clinical interpretation on a linear scale ranging from 0% (smallest ability) to 100% (largest ability) [36]. Expected responses, determined based on the patient and item locations, were compared to the responses actually reported to compute residual and fit statistics, which were then used to assess the scale’s unidimensionality [37]. A good fit of the data with the model affirms invariant locations along the continuum and indicates that the measure can be used to compare manual ability across patients and diagnoses.

### Item selection

From the experimental version of the questionnaire, the ABILHAND-HS was refined through successive analyses of 305 assessments with the goal of selecting items that define a unidimensional and clinically relevant scale of manual ability. P values < 0.05 were considered significant for each of the following analysis steps:

1. *Item-patient targeting*. Based on examination of patient distributions and item locations, items that showed a floor effect (too easy) or did not target the patients sample ability were removed.
2. *Rating scale*. Items with disordered thresholds and items with thresholds that were too narrow (<1.4 logits) or too wide (>5 logits) were removed before applying the rating scale model [38].
3. *Unidimensionality*. Only items that delineated a common manual ability construct according to the following four criteria were retained: (1) standardized residuals obtained over three class intervals had to be within ±2.5 with a non-significant χ^2^ [37]; (2) no observable major differential item functioning (DIF) [39], uniform or non-uniform, shown by a 2-way analysis of variance of the residuals with Bonferroni correction [40], according to gender (male vs. female), age (above vs. below the median age of 63 years), pathology (CTS vs. DRF vs. BTA vs. HWS), involved hand (dominant vs. non-dominant), level of education (basic vs. superior), and follow-up (0-3 months vs. 3–6 months vs. >6 months); (3) overall fit of the response set based on a non-significant item-trait interaction χ^2^ [37]; and (4) statistically similar patient locations, according to paired t-tests, calculated with items that loaded either positively or negatively on the first residuals principal component [41–43].
4. *Local independence*. When items were found to be querying redundant content [44], demonstrated by a residual correlation > 0.3, the item with the poorer fit statistic was deleted [45].
5. *Item redundancy*. To shorten the scale, when two or more items had similar locations on the continuum, the one with the best fit was retained.

### Scale reliability

The Person-Separation Index (PSI), i.e. the proportion of total variance (including error) that is attributed to patient location variance, was used to determine the ABILHAND-HS scale’s reliability and its degree of precision with the dataset, and thus how many statistically different ability strata can be distinguished along the scale [46].

### Construct validity

The construct validity of the ABILHAND-HS was examined with a comparison of means for associations with gender, involved hand, and diagnosis. The relationships of the ABILHAND-HS with age, the QuickDASH scale, the numerical pain scale, the SF-12 Physical Component Summary (PCS), and the SF-12 Mental Component Summary (MCS) were assessed with a correlation analysis.

Patient perceptions were compared between ABILHAND-HS and QuickDASH items by adding the six QuickDASH activity items to the anchored data matrix. The locations of similar items were then compared between the scales.

### Statistical analyses

Statistical analyses were completed in IBM SPSS Statistics for Windows, version 25 (IBM Corp., Armonk, N.Y., USA). Data normality was verified for statistical tests using the Shapiro-Wilk test and Q-Q plots. Parametric tests were used for normal data and continuous variables, non-parametric tests for non-normal data and ordinal variables. A Mann-Whitney u-test (two-tailed) was used for gender differences, an independent-samples t-test (two-tailed) for association with the involved hand, and an analysis of variance for diagnosis. Pearson correlation coefficient was calculated for association with age, while relationships with the QuickDASH scale, the numerical pain scale, the SF-12 Physical Component Summary (PCS), and the SF-12 Mental Component Summary (MCS) were assessed with Spearman correlation coefficients. P values < 0.05 were considered significant. Mean values are reported with standard deviations (SD). Chi-square and t values are reported with degrees of freedom (df).

## Results

### Item selection for the ABILHAND-HS scale

Successive analyses led to the selection of 23 items defining a unidimensional manual ability scale in HS. Of the 90 experimental items, 34 were removed because they were too easy (e.g. ‘Drinking a glass of water’), 3 items had too-narrow thresholds (e.g. ‘Using a touch screen’), 4 items were misfitting (e.g. ‘Carrying a shopping bag’), and 26 items had a location redundant with another better fitting item (e.g. ‘Peeling onions’ was deleted in favor of ‘Peeling potatoes with a knife’).

### Metric properties

The calibration obtained for the 23 mostly bimanual activities retained for ABILHAND-HS is reported in Table 2 in descending difficulty order. The standardized residuals obtained matched the expected standard normal distribution for items [mean (SD), −0.30 (0.99)] and for patients [0.31 (0.97)], indicating that the ABILHAND-HS scale is globally unidimensional. An invariant item location was obtained for more- and less-able patients as shown by a nonsignificant item-trait interaction (χ^2^ = 57.76, 46 df, p = 0.11). An invariant patient ability was obtained with items with different content as shown by a non significant t-test when using items that loaded positively or negatively on the first principal residual component (t = 1.24, 304 df, p = 0.22).

**Table 2.** Calibration of the 23 items of the ABILHAND-HS.

| Item | Bi-<br>manual | Difficulty logits<br>(centiles) | SE<br>logits | Residual<br>z | Fit<br>$\chi^2$ | P |
| --- | --- | --- | --- | --- | --- | --- |
| a. Doing push-ups | x | 3.54 (78) | 0.21 | 0.61 | 0.46 | 0.79 |
| b. Playing a racket sport | x | 2.30 (68) | 0.25 | 0.04 | 1.66 | 0.44 |
| c. Cutting a hedge | x | 2.00 (65) | 0.18 | 0.05 | 3.83 | 0.15 |
| d. Opening a screw-topped jar | x | 1.30 (59) | 0.12 | 0.48 | 6.19 | 0.05 |
| e. Applauding vigorously | x | 1.11 (58) | 0.16 | 2.09 | 2.00 | 0.37 |
| f. Lifting a full pan | x | 0.96 (57) | 0.12 | -0.53 | 5.69 | 0.06 |
| g. Wringing a towel | x | 0.86 (56) | 0.12 | -2.00 | 0.14 | 0.93 |
| h. Opening a can with a can opener | x | 0.76 (55) | 0.12 | -1.83 | 0.49 | 0.78 |
| i. Hammering a nail | x | 0.45 (52) | 0.16 | -0.70 | 5.31 | 0.07 |
| j. Shaking bed sheets | x | 0.13 (50) | 0.17 | -1.95 | 3.49 | 0.17 |
| k. Using a screwdriver | x | -0.05 (48) | 0.14 | -0.14 | 0.54 | 0.77 |
| l. Peeling potatoes with a knife | x | -0.16 (47) | 0.13 | -0.75 | 5.75 | 0.06 |
| m. Ironing | x | -0.38 (45) | 0.15 | -0.44 | 3.12 | 0.21 |
| n. Taking the cap off a bottle | x | -0.52 (44) | 0.13 | -0.32 | 2.75 | 0.25 |
| o. Cutting one's nails | x | -0.55 (44) | 0.13 | -0.05 | 0.33 | 0.85 |
| p. Shuffling and dealing cards | x | -0.77 (42) | 0.16 | -1.21 | 6.18 | 0.05 |
| q. Wiping windows |  | -0.77 (42) | 0.15 | 0.01 | 1.29 | 0.52 |
| r. Tying shoelaces | x | -0.96 (41) | 0.14 | -0.61 | 2.02 | 0.36 |
| s. Tearing open a pack of chips | x | -1.16 (39) | 0.15 | 1.70 | 1.31 | 0.52 |
| t. Fastening the zipper of a jacket | x | -1.45 (37) | 0.14 | 0.32 | 1.58 | 0.45 |
| u. Turning a car steering wheel | x | -1.69 (35) | 0.18 | -0.26 | 0.78 | 0.68 |
| v. Putting on gloves | x | -2.24 (30) | 0.19 | -0.80 | 1.99 | 0.37 |
| w. Spreading butter on a slice of bread | x | -2.68 (26) | 0.18 | -0.60 | 0.86 | 0.65 |

Analysis of DIF of the ABILHAND-HS with six criteria yielded only four instances of uniform DIF among the 23 items (Table 3). A small magnitude DIF was revealed among diagnoses (Fig 1) with no substantial impact on scale invariance, as evidenced by a good overall fit. Note that items were not specifically calibrated to the HWS group because of a limited sample size (n=23) [47]. No DIF was observed between the first and last assessments, showing satisfactory invariance to support the scale follow-up stability. Likewise, an intraclass correlation coefficient across the first and last assessments was 0.94, indicating excellent item-difficulty-hierarchy consistency and providing confidence for data pooling over different time points [48]. The PSI in this sample was equal to 0.90, indicating the distinguishability of four strata of manual ability [46].

**Table 3.**
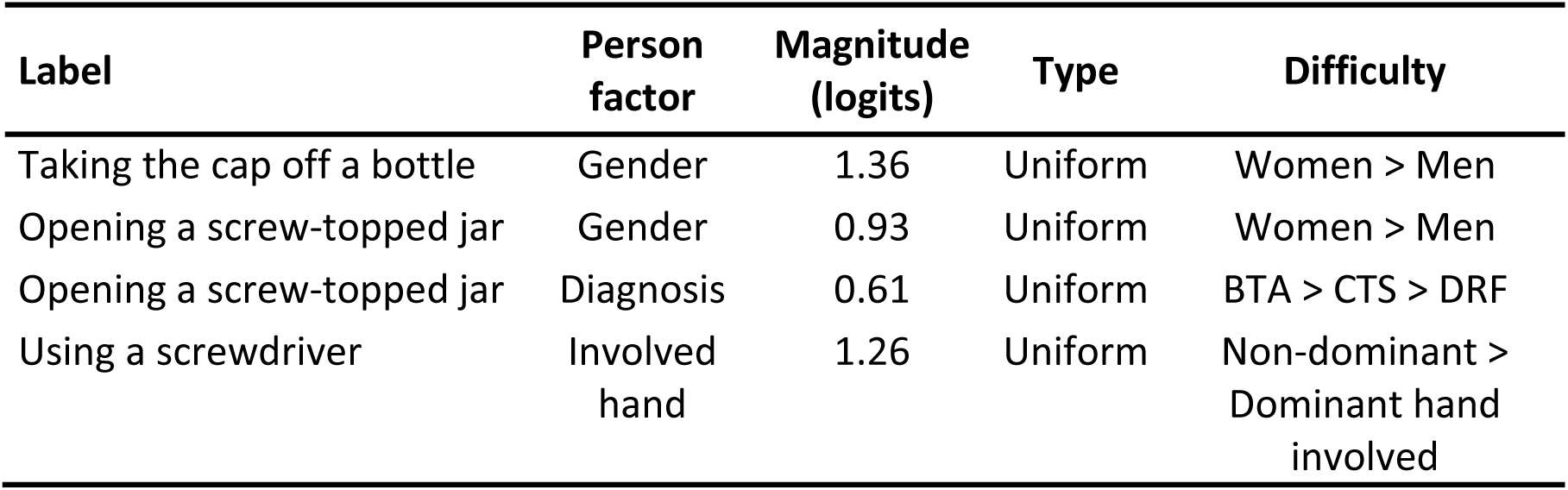
Differential item functioning (DIF) summary.

**Fig 1.**
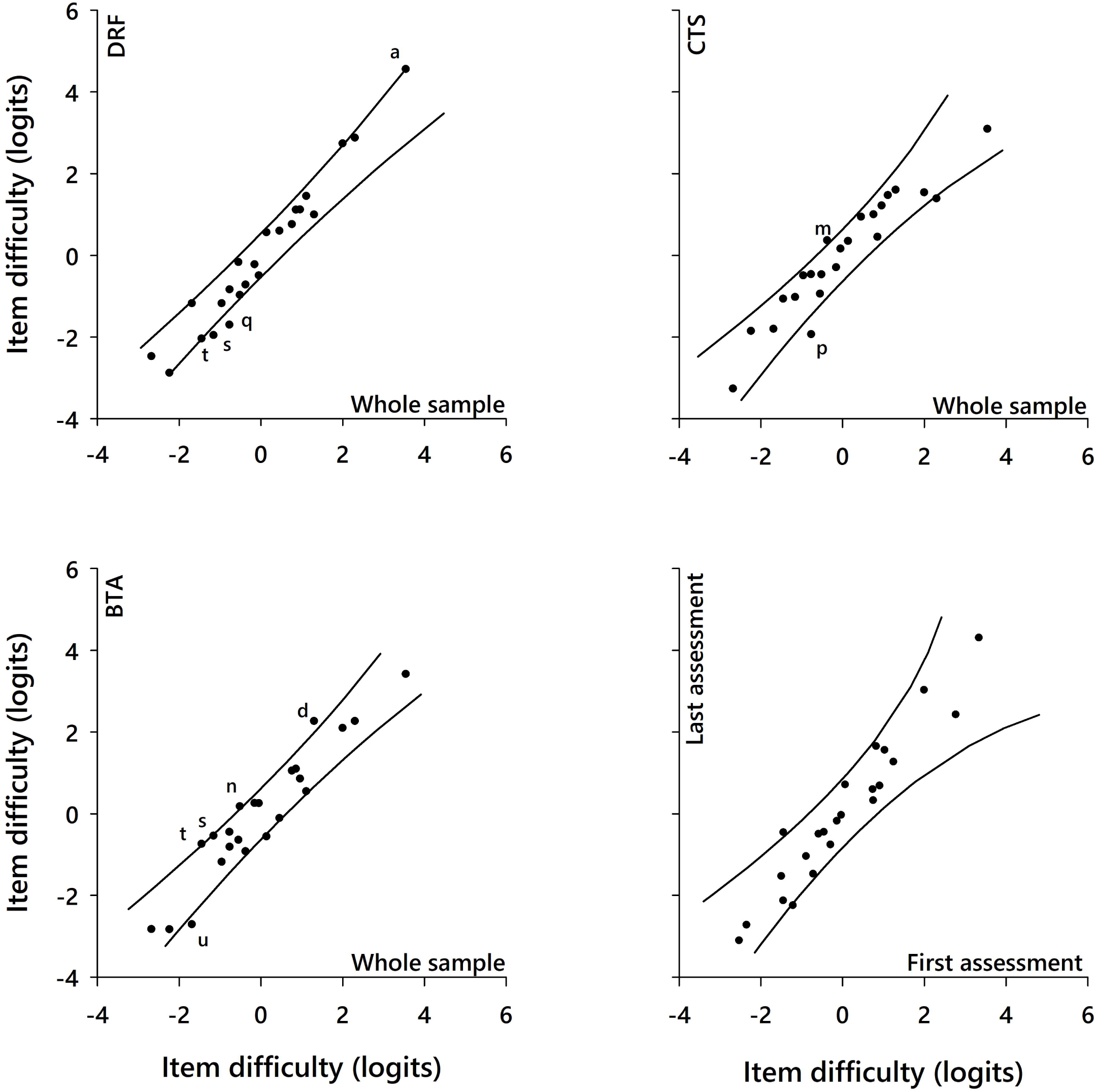
Differential item functioning (DIF) plots comparing the item difficulty hierarchy between subgroups. In each plot, the lines represent the 95% confidence interval of an ideal invariance between subgroups; the items are represented by the dots or by their letter if they display significant DIF. The most difficult items (dots) are plotted in the top right part of each plot. When comparing the item difficulty hierarchy between each diagnostic group relative to the whole sample, most of the ABILHAND-HS items lie within 95% confidence interval of the ideal invariance, indicating an invariant difficulty across diagnostic groups. When comparing the item difficulty hierarchy between the first and last assessment, all items fall within the 95% confidence interval of an ideal invariance, affirming invariance of item difficulties between the assessments.

### Scale description

The ABILHAND-HS structure and targeting of HS patients are illustrated in Fig 2, showing an average patients’ manual ability of 1.17 logits (SD = 1.85 logits; i.e. 58 (15) centiles). Twenty-four patients (7.9%) were able to perform all 23 activities easily, and were thus identified as extreme patients. Extreme patients tended to be younger men evaluated more than 6 months after treatment, and were more likely to have a CTS rather than a HWS. The three response categories were well distinguished in HS patients, with an inter-threshold distance of 2.93 logits (24 centiles), indicating that, regardless of patient ability, rating an item as ‘easy’ is about 20 (i.e. e^2.93^=18.7) times more difficult than rating it as ‘impossible’. Although the threshold distribution (range, - 4.15 to 5 logits) was well targeted to the range of patient abilities, the patients’ ability levels skewed high, indicating that the scale could measure patients that are more severely disabled than in this sample.

**Fig 2.**
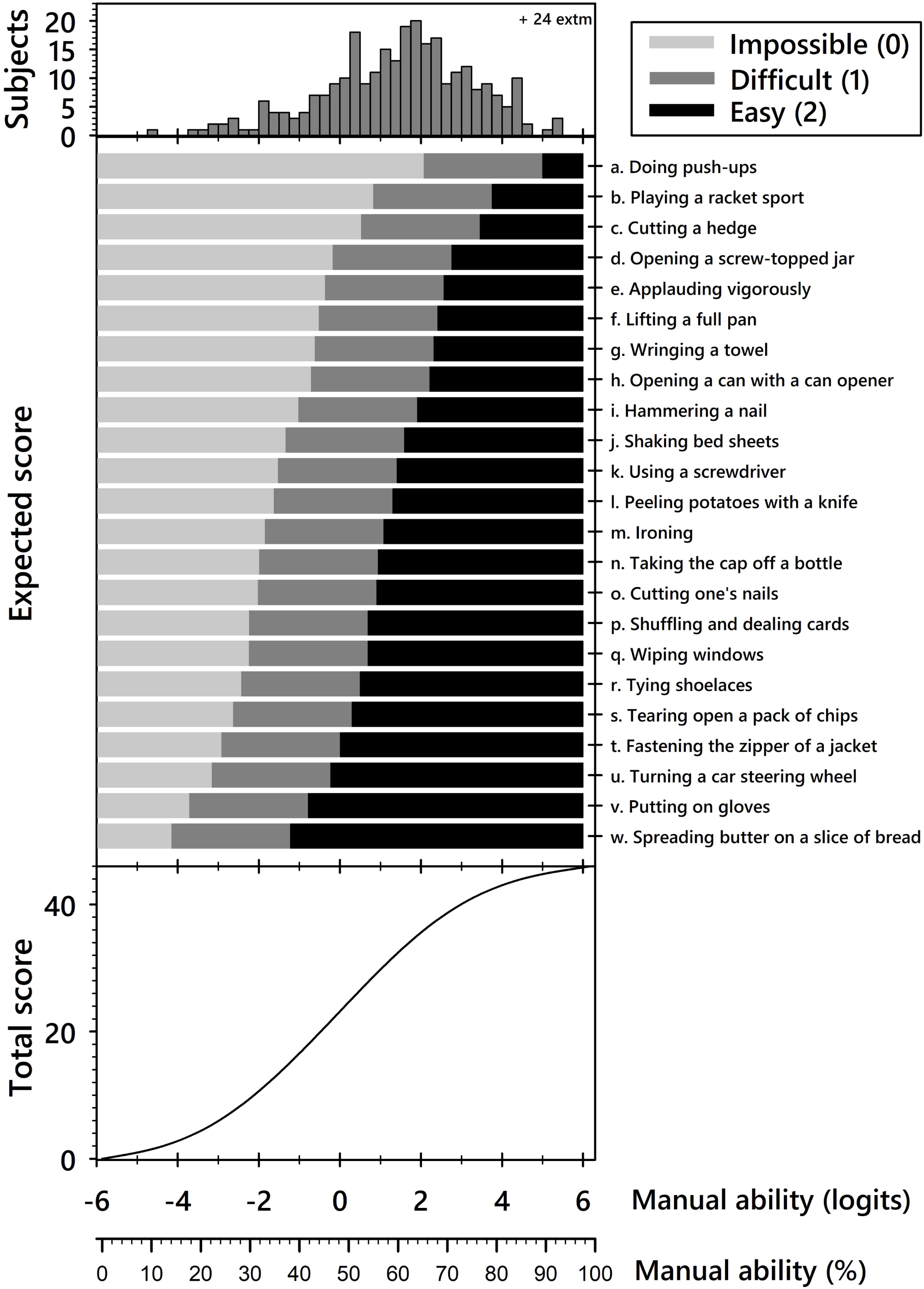
Structure of the ABILHAND-HS scale. Top: distribution of manual ability measures for the whole sample expressed in logits (log of the pass/fail probability ratio) and centiles (fraction of the measurement range). Twenty-four patients (7.9%) were able to perform all 23 activities easily, and were thus identified as extreme patients. None of the participants reported that they could not perform any of the 23 activities. Middle: most probable patient response to each item based on the patient manual ability and on the difficulty of the item’s response category. The average item difficulty was set to 0 logits and the items are ordered from most (top) to least (bottom) difficult. The distance between thresholds (middle bar) is constant for all items (2.93 logits or 24 centiles). A patient with a manual ability measure of 0 logits would be expected to perform the first 3 activities easily, to have some difficulty with the following 17 activities, and to be unable to perform the 3 most difficult activities. A patient with a measure of 2.1 logits should be able to perform all activities easily or with some difficulty. Bottom: conversion of ordinal raw scores into a linear continuum of manual ability for complete response sets. The raw scores ranged from 0 to 46 (sum of scores of 0–2 for 23 items). This curve is linear in its central (30^th^∼70^th^ percentile) range, with sigmoid flattening outside the central range, highlighting a non-linear relationship, especially at the extremities of the score range.

### Construct validity

ABILHAND-HS measures were normally distributed across the whole sample and subgroups, except for men (W = 0.97; 100 df; p = 0.038). An effect of gender on ABILHAND-HS manual ability measures was observed, with men [1.88 (2.39) logits; median 1.74 logits] reporting a significantly higher mean manual ability than women [1.32 (1.93) logits; median 1.4 logits; U = 8642; p = 0.026]. Manual ability was not found to be significantly associated with age (R = −0.04; p = 0.47), the hand involved (t = 0.96; 303 df; p = 0.37), or the patient’s diagnosis (F = 1.92; 3 df; p = 0.12). Although variance across diagnosis groups was not significant, we did observe a broad spectrum of manual ability. Patients with CTS reported the highest manual ability [1.9 (2.0) logits], followed by patients with BTA [1.5 (2.3) logits], DRF [1.4 (2.1) logits], and HWS [0.9 (1.8) logits].

The relationships between ABILHAND-HS measures and scores obtained with other instruments are shown in Fig 3. Briefly, ABILHAND-HS correlated strongly with QuickDASH scores, moderately with SF-12 PCS scores and pain scale scores, and weakly with SF-12 MCS scores. We observed substantial similarity with respect to manual ability scale locations between the ABLHAND-HS and QuickDASH activity items (Fig 4).

**Fig 3.**
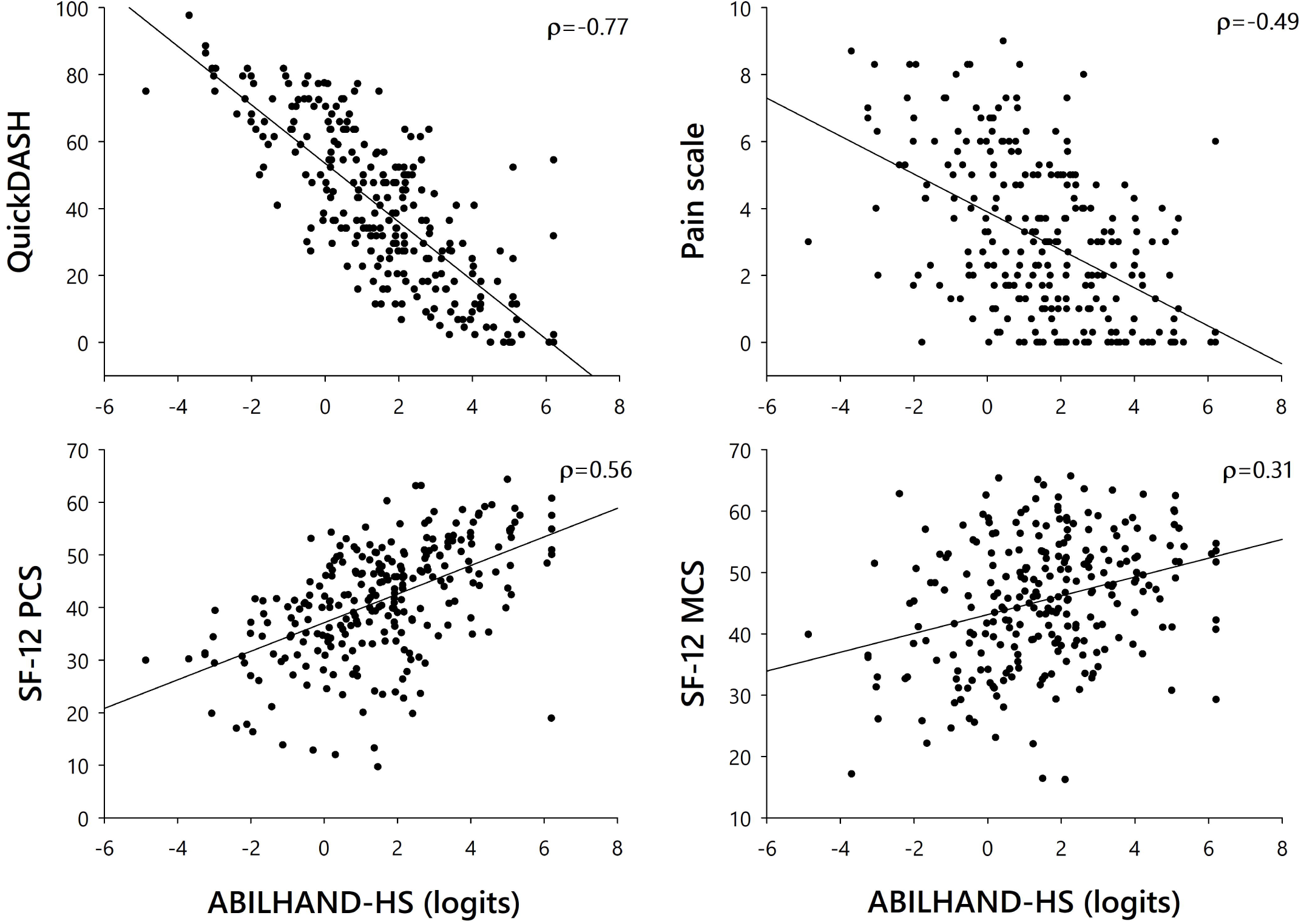
Correlations of ABILHAND-HS scores with QuickDASH, PCS, MCS, and numerical pain scale scores. Spearman correlation coefficients are indicated in the top right of each graph. All correlations were statistically significant (p < 0.001).

**Fig 4.**
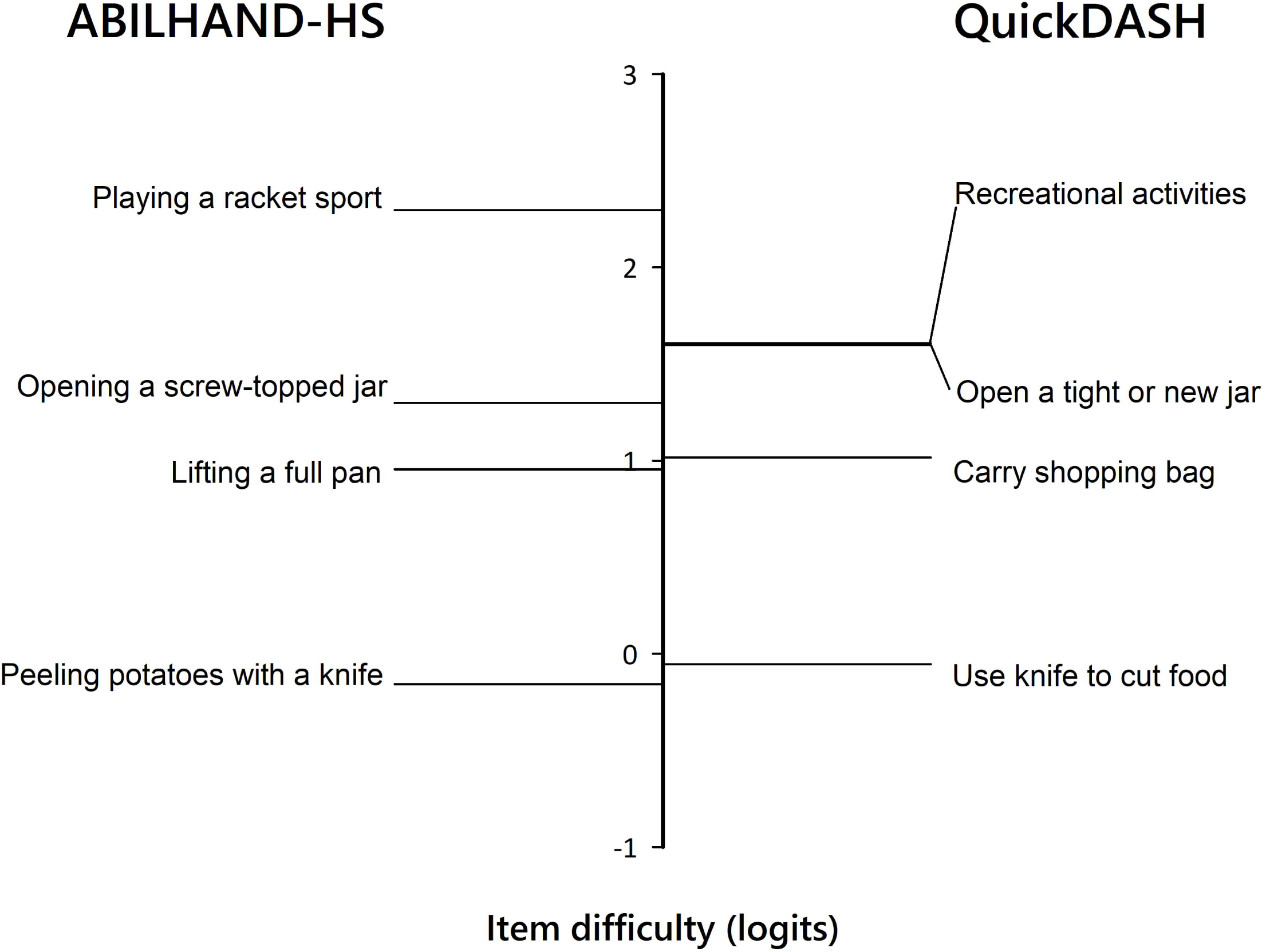
Comparison of difficulty levels (vertical axis) between similar items of the ABILHAND-HS (left) and QuickDASH (right) scales. QuickDASH item responses were added to the anchored data matrix of ABILHAND-HS responses to equate both measures.

## Discussion

Here, we report the adaptation and validation of an ABILHAND-HS questionnaire for use with HS patients. Impairments present in our study cohort included weakness (e.g. following DRF), loss of sensation (e.g. in CTS), and stiffness (e.g. in BTA), with some patients presenting with a combination of these impairments. The ABILHAND-HS was constructed to measure manual ability on a common, linear, and unidimensional scale wherein the 23 activities retained delineate an invariant item difficulty hierarchy independent of patient diagnosis. All ABILHAND-HS activities with the exception of one involve both hands and, consistent with our clinical experience, the most difficult ones require high levels of force (e.g. ‘doing push-ups’ loads the wrist in extension). Of the experimental 90 items, those that could be interpreted in different ways, for instance using the injured or uninjured hand, were misfitting and thus omitted (e.g. ‘carrying a shopping bag’). The sample size was adequate for the statistical interpretation of fit statistics [33], and was within the same range of studies dealing with the development of outcome measures [14,15,19,31]. The fit statistics for the 23 retained items support the item hierarchy invariance across the latent trait [49]. A few instances of minor DIF were retained to maintain the scale’s construct validity [50]. The resulting scale is well targeted to the studied HS population, despite a small persistent ceiling effect, most likely due to missing responses for the most difficult activities. This observation of apparent ceiling effect involves 7.9% (24/305) of the records, which is well below the maximum recommended allowance of 15% [51].

Although reliability indices should be compared with caution across potentially different study conditions, it is noteworthy that the PSI obtained for the ABILHAND-HS (0.90) was higher than prior values obtained for the activities subscales of the PRWE [52] (0.78 and 0.81 for the usual and specific activities subscales, respectively, in DRF patients), for the Patient-Rated Wrist and Hand Evaluation [53] (0.83 in HS patients), for the QuickDASH scale [54] (0.84 in patients with various upper limb dysfunctions) and for the Manual Ability Measure [55] (MAM-16; 0.83 for HS patients), while being equal to that for the DASH manual functioning subscale [18]. PSI values reflect sensitivity to clinical evolution over time, with greater values indicating a greater number of distinguishable ability strata. We obtained person separation among patients using three response levels (impossible, difficult, and easy), consistent with previous studies showing patients unable to discriminate more than three levels of difficulty (ABILHAND [8], DASH activity items [18] and QuickDASH activity items [54]).

Accurate communication of scale administration instructions is critical for targeting patient manual ability as defined by the ABILHAND-HS. Generally, patients focus on their ability to perform the queried activities with their injured hand; likewise, the PRWE explores use of the affected hand explicitly [15]. The ABILHAND-HS, like the QuickDASH, is oriented towards real daily life behaviors and is intended to be independent of the limb(s) or strategy used and unbiased by activities that are never performed with the affected hand or avoided during recovery [8]. Our findings of stable item calibrations and lack of DIF across the assessments indicate that the ABILHAND-HS can be used confidently to assess the patient recovery at different time points during follow-up. Moreover, the stability of items hierarchy between the first and last evaluation indicate that the results were not influenced by the method of administration (interview with the investigator versus self-reported).

ABILHAND-HS construct validation results fit well with our clinical observations. The patients with the highest manual ability scores on the ABILHAND-HS also had the highest SF-12 PCS and SF-12 MCS scores as well as the lowest QuickDASH and numerical pain scale scores, which was also observed in other validation studies [15,19]. Correlations of ABILHAND-HS with other instruments, including the QuickDASH, SF-12 and a pain scale are also consistent with prior findings suggesting that generic instruments are less sensitive than specific ones [56]. The present ABILHAND-HS manual ability scores were not related significantly to age, consistent with other versions of the ABILHAND [8,26,29]. Our findings of a small, but significant gender effect, with men tending to report a higher manual ability than women (mean difference, 0.56 logits), varied across HS diagnoses but, generally, were consistent with previous reports in patients with DRFs and wrist arthrodesis [57–59]. A possible explanation is that manual ability is related to grip strength, which is more important in men compared to women [8,60]. The construct validity of the ABILHAND-HS was further supported by our confirmation of a similar item difficulty hierarchy for QuickDASH items in our patients sample. Notably, those ABILHAND-HS activities that require a great amount of force (e.g. ‘Opening a screw-topped jar’) have been reported to likewise be among the most difficult items in the DASH and QuickDASH [18,54] and in the MAM-16 [55].

The ABILHAND-HS, developed using Rasch methodology, has several advantages over questionnaires developed using classical test theory. These are summarized in Table 4. Firstly, the ABILHAND-HS can tolerate missing responses, which enables it to remain valid even in patients who scarcely perform some of the queried activities. Secondly, the ability to analyze response patterns can identify those patients whose responses do not fit the model due to random or careless answers, a particular injury or comorbidities. Finally, the high precision of the ABILHAND-HS items minimizes the need for interpretation, thereby allowing more reliable comparisons between patients (e.g. recreational activities involving force or impact are broken down into the items ‘doing push-ups’, ‘practicing a racket sport’).

**Table 4.** Pros and cons of the ABILHAND-HS.

|  | Feature | Benefit |
| --- | --- | --- |
| **Pros** | Unidimensional and linear scale | Quantitative comparisons of manual ability can be made between patients, between treatments and along follow up |
|  | Invariant scale calibration validated in four diagnostic groups | Unbiased comparisons of manual ability can be made within and between clinical subgroups (different HS diagnostics, stage of recovery) |
|  | Precise item definition | Inaccurate responses and guessing are avoided |
|  | Amenable to incomplete responses | The test is specific to activities really performed by the patient |
|  | Capacity to analyze response patterns | The test can identify unexpected patient responses linked to patient specific behaviors, random or careless answers or comorbidities |
|  | Precision of the measure | The standard error of measurement is specific to each patient measure allowing statistical assessment during follow-up |
|  | Feature | Compensation |
| **Cons** | Yet another test for hand surgery outcomes evaluation | Takes five minutes to complete |
|  | Complex statistical background | Necessary for the analyst, but not for routine clinical use |
|  | Use of dedicated computer programs | A free web service (www.rehab-scales.org) can be used to interpret patient responses |
|  | Ceiling effect of 7.9% of records | Well below the maximum recommended allowance of 15% |

Limitations of this research include a sample of patients with hand and wrist disabilities from one hand surgery outpatient clinic. The unbalanced diagnostic groups and genders might have influenced the item calibrations. However, the gender distribution is similar in studies involving DRF and CTS [15,16,31] and the DIF analysis allowed us to select items where the diagnosis and gender effects were absent. Future studies with a larger sample size should confirm or refine our findings. Furthermore, the way participants responded to the 90 items in the questionnaire may not be the same as responses to the final 23-item instrument. One limitation to the availability of Rasch model-based questionnaires is that they have a complex statistical background and require the use of dedicated computer programs that are not easy to learn and implement. To facilitate and spread the use of the ABILHAND-HS, a website (www.rehab-scales.org) developed by Université catholique de Louvain and Arsalis, a spin-off of the ABILHAND authors’ laboratory, can be used to convert the questionnaire raw scores into manual ability measures. The web service is free-to-use for daily practice in clinical and research applications although a license is required for commercial applications and for clinical trials.

## Conclusion

ABILHAND-HS was demonstrated to be a successful adaptation for application in HS patients. The resulting scale was shown to be a valid, patient-oriented, clinically meaningful and precise instrument. It targets commonly performed manual activities and allows stable and linear measurement of manual ability over multiple time points in patients treated for DRF, BTA, CTS, or HWS. The scale reveals unexpected responses that may provide clues regarding the patient’s clinical state, as summarized at www.rehab-scales.org. The questionnaire is available online, and the web service is free-to-use for daily practice in clinical and research applications although a license is required for commercial applications and for clinical trials. Future research should include more patients with HWS, as well as other diagnoses such as tendinopathies, ligamentous injuries and complex hand injuries, and an assessment of scale responsiveness.

## Data Availability

The data in the study is based on patient material, and since there are many individual variables, there is a possibility that patients might be identified due to their comorbidities, patient characteristics and time of encounter. The study participants have not provided consent to publicly share the individual level data underlying this study. To ensure adherence to guidelines for the protection of data derived from human subjects, the patients records of this study as well as individual responses to the questionnaire can be obtained upon request to the ethical committee of Cliniques Universitaires Saint-Luc, with contact information, after identification of the applicant and justification of the demand.

## Supporting information

**S1 Text. Common versus specific scales**.

**S2 Text. The ABILHAND-HS questionnaire (English version) instructions and scoring sheets**. The scoring sheets are available in 10 different randomizations, as found on www.rehab-scales.org.

**S3 Text. The ABILHAND-HS questionnaire (French version) instructions and scoring sheets**. The scoring sheets are available in 10 different randomizations, as found on www.rehab-scales.org.

**S4 Appendix. The experimental 90-item ABILHAND questionnaire**.

