## Supplemental Text 1 for "Manual ability in hand surgery patients: validation of the ABILHAND scale in four diagnostic groups"

Supporting information

### **Common versus specific scales**

The range of measurement and the precision of a scale developed for the heterogenous patient group (CTS, DRF, BTA, and HWS patients) were compared to those obtained for diagnosis-specific scales constructed using the same methodology. There was not an HWS-specific scale developed due to the limited HWS sample size.

As shown in Figure 1, comparisons of the difficulty ratings of each of the 23 ABILHAND-HS items for the whole sample versus the difficulty ratings obtained by specific diagnostic groups (CTS, DRF, and BTA) demonstrated an absence of major DIF. Hence, we observed functional equivalence of the items with respect to item-difficulty hierarchy, indicating that the ABILHAND-HS should be applicable with invariance across patients with different HS diagnoses.

The ranges of measurement and precision levels obtained with the ABILHAND-HS (common scale) versus those obtained with ABILHAND-HS scales developed specifically for particular diagnostic groups (specific scales) are reported in S1 Table. The median error, range of measurement, and PSI values obtained for each of the diagnosis-specific scales were similar to those obtained for the common scale though there were fewer patients with extreme scores when the whole cohort was analyzed together, confirming that the scale benefits from a larger number of responses from a clinically heterogenous cohort. The correlations between patient abilities obtained on the common and specific scales were very high, further supporting the inferred equivalence of both types of scales.

**S1 Table.** **Correlation of performance of the diagnostic group-specific scales with that of the common ABILHAND-HS scale.**

| ***Diagnostic group*** | ***Specific scale*** | | | |  | ***Common scale*** | | | ***Correlation***  ***coefficient***  ***R*** |
| --- | --- | --- | --- | --- | --- | --- | --- | --- | --- |
|  | ***Median SE,***  ***logits*** | ***Range, logits*** | ***Extreme subjects, %*** | ***PSI*** |  | ***Median SE,***  ***logits*** | ***Range, logits*** | ***Extreme subjects, %*** |  |
| DRF | 0.57 | 9.28 | 11.2 | 0.90 |  | 0.52 | 9.49 | 7.2 | 0.99 |
| CTS | 0.52 | 9.32 | 12.0 | 0.90 |  | 0.57 | 9.14 | 12 | 0.97 |
| BTA | 0.60 | 10.3 | 12.6 | 0.92 |  | 0.49 | 9.08 | 7 | 0.97 |

The ABILHAND-HS scale performed at least as well as scales specific to CTS, DRS, and BTA diagnoses, while allowing manual ability to be compared across HS diagnostic groups with a simple single scale. Although prior results concerning the applicability of a generic ABILHAND scale have been mixed [1,2], the present results can be explained by the greater homogeneity of our case mix compared to that of Arnould et al. [1] and by our selection of only those items with diagnosis-independent difficulty for inclusion in the ABILHAND-HS. The slight differences obtained with our diagnosis-specific scales relative to our more generic scale (retained as the ABILHAND-HS) also support the clinical validity of the ABILHAND-HS scale. For example, low-force-requirement activities (e.g. ‘Fastening the zipper of a jacket’) proved to be relatively easier for patients treated for a DRF than for BTA, for whom finger pinch movements are particularly painful.
