## Supplemental Text 2 for "Manual ability in hand surgery patients: validation of the ABILHAND scale in four diagnostic groups"

**Université catholique de Louvain	Haute Ecole Louvain en Hainaut**

Laboratory of Rehabilitation	Physical and Occupational Therapy Departments

and Physical medicine	Paramedical Category

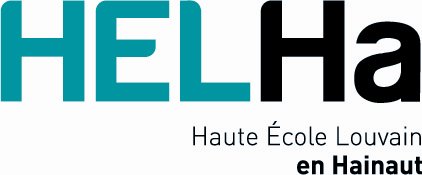

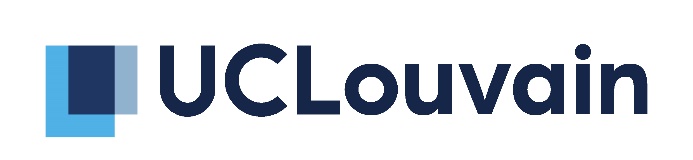

**________________________________________________________________________________________**

**Instructions for the ABILHAND-HS questionnaire**

### **The ABILHAND-HS questionnaire**

The ABILHAND-HS (Hand Surgery) questionnaire was developed with hand surgery patients as a measure of manual ability as perceived by the patient. It explores the most representative inventory of manual activities. Some items were selected from existing scales; others were devised to extend the range of activities. The original questionnaire was applied to a sample of rheumatoid arthritis patients (Arch Phys Med Rehabiil 1998; 79: 1038-42). The scale has since been validated in populations with rheumatoid arthritis (Ann Rheum Dis 2007; 66: 1098-1105), chronic stroke (Stroke 2001; 32: 1627-1634), pediatric cerebral palsy (Neurology 2004; 63: 1045-1052), systemic sclerosis (J Neurol Neurosurg Psychiatr 2010; 81: 506-512) and neuromuscular disease (J Neurol Neurosurg Psychiatr 2010; 81: 506-512). ABILHAND was originally developed using the Rasch measurement model. It allows to convert ordinal scores into linear measures located on a unidimensional scale.

### **Procedures**

The ABILHAND-HS questionnaire is administered on an interview basis (patients do not realize the activities). Patients are asked to estimate the ease or difficulty in performing each activity, when the activities are done:

- Without other technical or human help (even if the patient actually uses help in daily life)
- Irrespective of the limb(s) actually used to do the activity
- Whatever the strategy used (any compensation is allowed)

During the evaluation, a 3-level response scale is presented to the patients. Patients are asked to rate their perception on the response scale as either “Impossible”, “Difficult” or “Easy”. Activities not attempted in the last week are not scored and are entered as missing responses (tick the question mark). For any activity the four potential answers are:

- ***Impossible :*** the patient is unable to perform the activity without using any other help
- ***Difficult :*** the patient is able to perform the activity without any help but experiences some difficulty
- ***Easy :*** the patient is able to perform the activity without any help and experiences no difficulty
- ***Question mark:*** the patient cannot estimate the difficulty of the activity because he/she has never done the activity. Note that when a patient has never attempted the activity, the rater needs to make sure why it is so. If an activity was never attempted because it is impossible, then it must be scored as “Impossible” rather than “Question mark”.

The instructions are given to the patient only at the beginning of the test. Five items are used for training in order to help the patient in feeling each level of the rating scale and in using the whole amplitude of the response scale. The subsequent activities are neither preceded nor followed by any instruction. The examiner can repeat the instructions whenever the patient shows some hesitation in answering.

### **Activities order**

The activities of the ABILHAND-HS questionnaire are presented in a random order to avoid any systematic effect. Ten different random orders of presentation are used. The rated must select the next one of the 10 orders for each new assessment, no matter which patient is tested.

### **Package content**

- 1 instruction sheet
- Testing forms in 10 random orders (10 sheets)

**ABILHAND- Manual Ability Measure**

**English version**

Patient ______________________________________	Date ________________

|  | How DIFFICULT are the following activities? | Impossible (0) | Difficult (1) | Easy (2) | ? |
| --- | --- | --- | --- | --- | --- |
| 1. | Lifting a full pan |  |  |  |  |
| 2. | Ironing |  |  |  |  |
| 3. | Tearing open a pack of chips |  |  |  |  |
| 4. | Spreading butter on a slice of bread |  |  |  |  |
| 5. | Doing push-ups |  |  |  |  |
| 6. | Hammering a nail |  |  |  |  |
| 7. | Opening a can with a can opener |  |  |  |  |
| 8. | Using a screwdriver |  |  |  |  |
| 9. | Taking the cap off a bottle |  |  |  |  |
| 10. | Shaking bed sheets |  |  |  |  |
| 11. | Opening a screw-topped jar |  |  |  |  |
| 12. | Peeling potatoes with a knife |  |  |  |  |
| 13. | Wringing a towel |  |  |  |  |
| 14. | Cutting a hedge |  |  |  |  |
| 15. | Tying shoelaces |  |  |  |  |
| 16. | Playing a racket sport |  |  |  |  |
| 17. | Applauding vigorously |  |  |  |  |
| 18. | Turning a car steering wheel |  |  |  |  |
| 19. | Cutting one’s nails |  |  |  |  |
| 20. | Fastening the zipper of a jacket |  |  |  |  |
| 21. | Putting on gloves |  |  |  |  |
| 22. | Shuffling and dealing cards |  |  |  |  |
| 23. | Wiping windows |  |  |  |  |

Université catholique de Louvain & Haute Ecole Louvain en Hainaut ; [www.rehab-scales.org](http://www.rehab-scales.org)	Order 1

**ABILHAND- Manual Ability Measure**

**English version**

Patient ______________________________________	Date ________________

|  | How DIFFICULT are the following activities? | Impossible (0) | Difficult (1) | Easy (2) | ? |
| --- | --- | --- | --- | --- | --- |
| 1. | Opening a can with a can opener |  |  |  |  |
| 2. | Fastening the zipper of a jacket |  |  |  |  |
| 3. | Cutting one’s nails |  |  |  |  |
| 4. | Wiping windows |  |  |  |  |
| 5. | Turning a car steering wheel |  |  |  |  |
| 6. | Peeling potatoes with a knife |  |  |  |  |
| 7. | Using a screwdriver |  |  |  |  |
| 8. | Playing a racket sport |  |  |  |  |
| 9. | Opening a screw-topped jar |  |  |  |  |
| 10. | Shuffling and dealing cards |  |  |  |  |
| 11. | Doing push-ups |  |  |  |  |
| 12. | Ironing |  |  |  |  |
| 13. | Shaking bed sheets |  |  |  |  |
| 14. | Taking the cap off a bottle |  |  |  |  |
| 15. | Lifting a full pan |  |  |  |  |
| 16. | Hammering a nail |  |  |  |  |
| 17. | Cutting a hedge |  |  |  |  |
| 18. | Putting on gloves |  |  |  |  |
| 19. | Applauding vigorously |  |  |  |  |
| 20. | Tying shoelaces |  |  |  |  |
| 21. | Spreading butter on a slice of bread |  |  |  |  |
| 22. | Tearing open a pack of chips |  |  |  |  |
| 23. | Wringing a towel |  |  |  |  |

Université catholique de Louvain & Haute Ecole Louvain en Hainaut ; [www.rehab-scales.org](http://www.rehab-scales.org)	Order 2

**ABILHAND- Manual Ability Measure**

**English version**

Patient ______________________________________	Date ________________

|  | How DIFFICULT are the following activities? | Impossible (0) | Difficult (1) | Easy (2) | ? |
| --- | --- | --- | --- | --- | --- |
| 1. | Peeling potatoes with a knife |  |  |  |  |
| 2. | Wringing a towel |  |  |  |  |
| 3. | Tearing open a pack of chips |  |  |  |  |
| 4. | Taking the cap off a bottle |  |  |  |  |
| 5. | Using a screwdriver |  |  |  |  |
| 6. | Opening a screw-topped jar |  |  |  |  |
| 7. | Cutting one’s nails |  |  |  |  |
| 8. | Cutting a hedge |  |  |  |  |
| 9. | Turning a car steering wheel |  |  |  |  |
| 10. | Ironing |  |  |  |  |
| 11. | Opening a can with a can opener |  |  |  |  |
| 12. | Lifting a full pan |  |  |  |  |
| 13. | Applauding vigorously |  |  |  |  |
| 14. | Playing a racket sport |  |  |  |  |
| 15. | Wiping windows |  |  |  |  |
| 16. | Hammering a nail |  |  |  |  |
| 17. | Putting on gloves |  |  |  |  |
| 18. | Spreading butter on a slice of bread |  |  |  |  |
| 19. | Shuffling and dealing cards |  |  |  |  |
| 20. | Doing push-ups |  |  |  |  |
| 21. | Shaking bed sheets |  |  |  |  |
| 22. | Tying shoelaces |  |  |  |  |
| 23. | Fastening the zipper of a jacket |  |  |  |  |

Université catholique de Louvain & Haute Ecole Louvain en Hainaut ; [www.rehab-scales.org](http://www.rehab-scales.org)	Order 3

**ABILHAND- Manual Ability Measure**

**English version**

Patient ______________________________________	Date ________________

|  | How DIFFICULT are the following activities? | Impossible (0) | Difficult (1) | Easy (2) | ? |
| --- | --- | --- | --- | --- | --- |
| 1. | Hammering a nail |  |  |  |  |
| 2. | Using a screwdriver |  |  |  |  |
| 3. | Fastening the zipper of a jacket |  |  |  |  |
| 4. | Opening a can with a can opener |  |  |  |  |
| 5. | Doing push-ups |  |  |  |  |
| 6. | Playing a racket sport |  |  |  |  |
| 7. | Shuffling and dealing cards |  |  |  |  |
| 8. | Cutting a hedge |  |  |  |  |
| 9. | Putting on gloves |  |  |  |  |
| 10. | Cutting one’s nails |  |  |  |  |
| 11. | Spreading butter on a slice of bread |  |  |  |  |
| 12. | Tying shoelaces |  |  |  |  |
| 13. | Wringing a towel |  |  |  |  |
| 14. | Opening a screw-topped jar |  |  |  |  |
| 15. | Tearing open a pack of chips |  |  |  |  |
| 16. | Turning a car steering wheel |  |  |  |  |
| 17. | Peeling potatoes with a knife |  |  |  |  |
| 18. | Taking the cap off a bottle |  |  |  |  |
| 19. | Shaking bed sheets |  |  |  |  |
| 20. | Applauding vigorously |  |  |  |  |
| 21. | Lifting a full pan |  |  |  |  |
| 22. | Ironing |  |  |  |  |
| 23. | Wiping windows |  |  |  |  |

Université catholique de Louvain & Haute Ecole Louvain en Hainaut ; [www.rehab-scales.org](http://www.rehab-scales.org)	Order 4

**ABILHAND- Manual Ability Measure**

**English version**

Patient ______________________________________	Date ________________

|  | How DIFFICULT are the following activities? | Impossible (0) | Difficult (1) | Easy (2) | ? |
| --- | --- | --- | --- | --- | --- |
| 1. | Turning a car steering wheel |  |  |  |  |
| 2. | Tearing open a pack of chips |  |  |  |  |
| 3. | Spreading butter on a slice of bread |  |  |  |  |
| 4. | Tying shoelaces |  |  |  |  |
| 5. | Opening a screw-topped jar |  |  |  |  |
| 6. | Doing push-ups |  |  |  |  |
| 7. | Fastening the zipper of a jacket |  |  |  |  |
| 8. | Using a screwdriver |  |  |  |  |
| 9. | Cutting a hedge |  |  |  |  |
| 10. | Applauding vigorously |  |  |  |  |
| 11. | Wiping windows |  |  |  |  |
| 12. | Opening a can with a can opener |  |  |  |  |
| 13. | Wringing a towel |  |  |  |  |
| 14. | Ironing |  |  |  |  |
| 15. | Hammering a nail |  |  |  |  |
| 16. | Peeling potatoes with a knife |  |  |  |  |
| 17. | Playing a racket sport |  |  |  |  |
| 18. | Cutting one’s nails |  |  |  |  |
| 19. | Lifting a full pan |  |  |  |  |
| 20. | Shuffling and dealing cards |  |  |  |  |
| 21. | Taking the cap off a bottle |  |  |  |  |
| 22. | Shaking bed sheets |  |  |  |  |
| 23. | Putting on gloves |  |  |  |  |

Université catholique de Louvain & Haute Ecole Louvain en Hainaut ; [www.rehab-scales.org](http://www.rehab-scales.org)	Order 5

**ABILHAND- Manual Ability Measure**

**English version**

Patient ______________________________________	Date ________________

|  | How DIFFICULT are the following activities? | Impossible (0) | Difficult (1) | Easy (2) | ? |
| --- | --- | --- | --- | --- | --- |
| 1. | Cutting a hedge |  |  |  |  |
| 2. | Wringing a towel |  |  |  |  |
| 3. | Opening a can with a can opener |  |  |  |  |
| 4. | Shuffling and dealing cards |  |  |  |  |
| 5. | Lifting a full pan |  |  |  |  |
| 6. | Tearing open a pack of chips |  |  |  |  |
| 7. | Fastening the zipper of a jacket |  |  |  |  |
| 8. | Peeling potatoes with a knife |  |  |  |  |
| 9. | Shaking bed sheets |  |  |  |  |
| 10. | Doing push-ups |  |  |  |  |
| 11. | Applauding vigorously |  |  |  |  |
| 12. | Tying shoelaces |  |  |  |  |
| 13. | Using a screwdriver |  |  |  |  |
| 14. | Spreading butter on a slice of bread |  |  |  |  |
| 15. | Opening a screw-topped jar |  |  |  |  |
| 16. | Putting on gloves |  |  |  |  |
| 17. | Turning a car steering wheel |  |  |  |  |
| 18. | Playing a racket sport |  |  |  |  |
| 19. | Hammering a nail |  |  |  |  |
| 20. | Wiping windows |  |  |  |  |
| 21. | Ironing |  |  |  |  |
| 22. | Cutting one’s nails |  |  |  |  |
| 23. | Taking the cap off a bottle |  |  |  |  |

Université catholique de Louvain & Haute Ecole Louvain en Hainaut ; [www.rehab-scales.org](http://www.rehab-scales.org)	Order 6

**ABILHAND- Manual Ability Measure**

**English version**

Patient ______________________________________	Date ________________

|  | How DIFFICULT are the following activities? | Impossible (0) | Difficult (1) | Easy (2) | ? |
| --- | --- | --- | --- | --- | --- |
| 1. | Shuffling and dealing cards |  |  |  |  |
| 2. | Wiping windows |  |  |  |  |
| 3. | Hammering a nail |  |  |  |  |
| 4. | Tearing open a pack of chips |  |  |  |  |
| 5. | Turning a car steering wheel |  |  |  |  |
| 6. | Opening a can with a can opener |  |  |  |  |
| 7. | Putting on gloves |  |  |  |  |
| 8. | Shaking bed sheets |  |  |  |  |
| 9. | Applauding vigorously |  |  |  |  |
| 10. | Cutting one’s nails |  |  |  |  |
| 11. | Ironing |  |  |  |  |
| 12. | Wringing a towel |  |  |  |  |
| 13. | Opening a screw-topped jar |  |  |  |  |
| 14. | Taking the cap off a bottle |  |  |  |  |
| 15. | Fastening the zipper of a jacket |  |  |  |  |
| 16. | Lifting a full pan |  |  |  |  |
| 17. | Cutting a hedge |  |  |  |  |
| 18. | Using a screwdriver |  |  |  |  |
| 19. | Playing a racket sport |  |  |  |  |
| 20. | Tying shoelaces |  |  |  |  |
| 21. | Peeling potatoes with a knife |  |  |  |  |
| 22. | Doing push-ups |  |  |  |  |
| 23. | Spreading butter on a slice of bread |  |  |  |  |

Université catholique de Louvain & Haute Ecole Louvain en Hainaut ; [www.rehab-scales.org](http://www.rehab-scales.org)	Order 7

**ABILHAND- Manual Ability Measure**

**English version**

Patient ______________________________________	Date ________________

|  | How DIFFICULT are the following activities? | Impossible (0) | Difficult (1) | Easy (2) | ? |
| --- | --- | --- | --- | --- | --- |
| 1. | Using a screwdriver |  |  |  |  |
| 2. | Turning a car steering wheel |  |  |  |  |
| 3. | Cutting one’s nails |  |  |  |  |
| 4. | Taking the cap off a bottle |  |  |  |  |
| 5. | Spreading butter on a slice of bread |  |  |  |  |
| 6. | Ironing |  |  |  |  |
| 7. | Shaking bed sheets |  |  |  |  |
| 8. | Tearing open a pack of chips |  |  |  |  |
| 9. | Hammering a nail |  |  |  |  |
| 10. | Lifting a full pan |  |  |  |  |
| 11. | Opening a screw-topped jar |  |  |  |  |
| 12. | Shuffling and dealing cards |  |  |  |  |
| 13. | Wringing a towel |  |  |  |  |
| 14. | Peeling potatoes with a knife |  |  |  |  |
| 15. | Fastening the zipper of a jacket |  |  |  |  |
| 16. | Putting on gloves |  |  |  |  |
| 17. | Applauding vigorously |  |  |  |  |
| 18. | Wiping windows |  |  |  |  |
| 19. | Tying shoelaces |  |  |  |  |
| 20. | Cutting a hedge |  |  |  |  |
| 21. | Playing a racket sport |  |  |  |  |
| 22. | Doing push-ups |  |  |  |  |
| 23. | Opening a can with a can opener |  |  |  |  |

Université catholique de Louvain & Haute Ecole Louvain en Hainaut ; [www.rehab-scales.org](http://www.rehab-scales.org)	Order 8

**ABILHAND- Manual Ability Measure**

**English version**

Patient ______________________________________	Date ________________

|  | How DIFFICULT are the following activities? | Impossible (0) | Difficult (1) | Easy (2) | ? |
| --- | --- | --- | --- | --- | --- |
| 1. | Tying shoelaces |  |  |  |  |
| 2. | Applauding vigorously |  |  |  |  |
| 3. | Hammering a nail |  |  |  |  |
| 4. | Cutting a hedge |  |  |  |  |
| 5. | Doing push-ups |  |  |  |  |
| 6. | Putting on gloves |  |  |  |  |
| 7. | Peeling potatoes with a knife |  |  |  |  |
| 8. | Lifting a full pan |  |  |  |  |
| 9. | Wringing a towel |  |  |  |  |
| 10. | Shaking bed sheets |  |  |  |  |
| 11. | Opening a screw-topped jar |  |  |  |  |
| 12. | Tearing open a pack of chips |  |  |  |  |
| 13. | Spreading butter on a slice of bread |  |  |  |  |
| 14. | Ironing |  |  |  |  |
| 15. | Taking the cap off a bottle |  |  |  |  |
| 16. | Fastening the zipper of a jacket |  |  |  |  |
| 17. | Wiping windows |  |  |  |  |
| 18. | Using a screwdriver |  |  |  |  |
| 19. | Turning a car steering wheel |  |  |  |  |
| 20. | Playing a racket sport |  |  |  |  |
| 21. | Opening a can with a can opener |  |  |  |  |
| 22. | Shuffling and dealing cards |  |  |  |  |
| 23. | Cutting one’s nails |  |  |  |  |

Université catholique de Louvain & Haute Ecole Louvain en Hainaut ; [www.rehab-scales.org](http://www.rehab-scales.org)	Order 9

**ABILHAND- Manual Ability Measure**

**English version**

Patient ______________________________________	Date ________________

|  | How DIFFICULT are the following activities? | Impossible (0) | Difficult (1) | Easy (2) | ? |
| --- | --- | --- | --- | --- | --- |
| 1. | Hammering a nail |  |  |  |  |
| 2. | Wiping windows |  |  |  |  |
| 3. | Applauding vigorously |  |  |  |  |
| 4. | Tying shoelaces |  |  |  |  |
| 5. | Opening a can with a can opener |  |  |  |  |
| 6. | Doing push-ups |  |  |  |  |
| 7. | Using a screwdriver |  |  |  |  |
| 8. | Shuffling and dealing cards |  |  |  |  |
| 9. | Fastening the zipper of a jacket |  |  |  |  |
| 10. | Putting on gloves |  |  |  |  |
| 11. | Shaking bed sheets |  |  |  |  |
| 12. | Playing a racket sport |  |  |  |  |
| 13. | Opening a screw-topped jar |  |  |  |  |
| 14. | Spreading butter on a slice of bread |  |  |  |  |
| 15. | Cutting a hedge |  |  |  |  |
| 16. | Cutting one’s nails |  |  |  |  |
| 17. | Lifting a full pan |  |  |  |  |
| 18. | Tearing open a pack of chips |  |  |  |  |
| 19. | Peeling potatoes with a knife |  |  |  |  |
| 20. | Ironing |  |  |  |  |
| 21. | Turning a car steering wheel |  |  |  |  |
| 22. | Wringing a towel |  |  |  |  |
| 23. | Taking the cap off a bottle |  |  |  |  |

Université catholique de Louvain & Haute Ecole Louvain en Hainaut ; [www.rehab-scales.org](http://www.rehab-scales.org)	Order 10
