## Supplemental Text 3 for "Manual ability in hand surgery patients: validation of the ABILHAND scale in four diagnostic groups"

**Université catholique de Louvain	Haute Ecole Louvain en Hainaut**

Laboratory of Rehabilitation	Physical and Occupational Therapy Departments

and Physical medicine	Paramedical Category

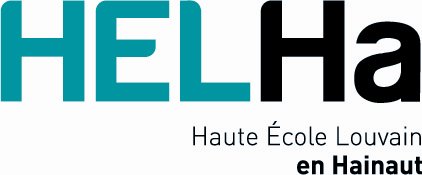

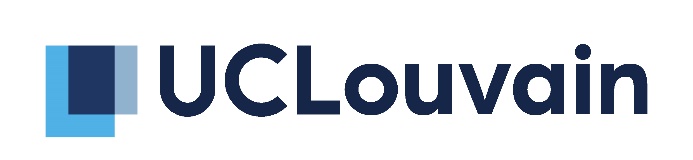

**_____________________________________________________________________________________________**

**Instructions pour le questionnaire ABILHAND-HS**

### **Le questionnaire ABILHAND-HS**

Le questionnaire ABILHAND-HS (Hand Surgery) a été développé auprès de patients en chirurgie de la main afin de mesurer l’habileté manuelle perçue par le patient. Il explore les activités manuelles les plus représentatives de la vie quotidienne. Certains items ont été sélectionnés à partir d’échelles existantes, d’autres ont été rajoutés afin d’étendre la gamme des activités. La première application du questionnaire dans un échantillon de patients atteints d’arthrite rhumatoïde (Arch Phys Med Rehabiil 1998; 79: 1038-42) a montré que les items définissent une échelle d’habileté manuelle valide. L’échelle a depuis été validé pour des patients atteints de polyarthrite rhumatoïde (Ann Rheum Dis 2007; 66: 1098-1105), d’hémiplégie chronique (Stroke 2001; 32: 1627-1634), d’infirmité motrice cérébrale (Neurology 2004; 63: 1045-1052), de sclérodermie (J Neurol Neurosurg Psychiatr 2010; 81: 506-512), et de maladies neuromusculaires (J Neurol Neurosurg Psychiatr 2010; 81: 506-512). ABILHAND a été développé en utilisant le modèle de Rasch. Ce modèle permet de convertir des scores ordinaux en mesures linéaires localisées sur une échelle unidimensionnelle.

### **Procédures**

Le questionnaire ABILHAND-HS est administré sous forme d’interview (les patients ne réalisent donc pas les activités). Les patients doivent estimer la difficulté de chaque activité lorsque les activités sont réalisées :

- Sans aide technique ni humaine (même si le patient utilise habituellement une aide dans la vie quotidienne)
- Quel(s) que soi(en)t le(s) membre(s) utilisé(s) pour réaliser l’activité
- Quelle que soit la stratégie utilisée (toutes les compensations sont autorisées)

Une échelle à 3 catégories de réponses est présentée. Les patients estiment la difficulté de chaque activité comme "Impossible", "Difficile" ou "Facile". Les activités qui n’ont pas été réalisées durant la dernière semaine ne sont pas cotées et sont considérées comme des réponses manquantes (cocher le point d’interrogation). Pour chaque activité, 4 réponses sont possibles :

- ***Impossible :*** le patient est incapable de réaliser l’activité sans l’utilisation d’une aide extérieure
- ***Difficile :*** le patient est capable de réaliser l’activité sans aide mais éprouve néanmoins quelques difficultés
- ***Facile :*** le patient est capable de réaliser l’activité sans aide et n’éprouve aucune difficulté à la réaliser
- ***Point d’interrogation :*** le patient incapable d’estimer la difficulté de l’activité parce qu’il/elle n’a pas réalisé l’activité. L’examinateur doit impérativement s’assurer de la raison pour laquelle le patient n’a pas réalisé l’activité. Si l’activité n’a jamais été réalisée parce qu’elle est impossible, vous devez cocher "Impossible" plutôt que "Point d’interrogation".

Les instructions sont données aux patients uniquement au début du test. Cinq items sont utilisés en guise d’entraînement pour aider le patient à comprendre chacune des catégories de l’échelle et afin d’utiliser toute l’amplitude de l’échelle de réponses. Aucune autre instruction ultérieure n’est nécessaire. L’examinateur peut toutefois répéter les instructions si le patient montre quelques hésitations dans ses réponses.

### **Ordre des activités**

Les acticités du questionnaire ABILHAND-HS sont présentées dans un ordre aléatoire afin d’éviter un biais systématique. Il existe dix ordres de présentation aléatoires différents. L’examinateur doit sélectionner, à chaque nouvelle évaluation, l’ordre suivant le dernier utilisé et ce quel que soit le patient testé.

### **Contenu du document**

- 1 feuille d’instruction
- Les formulaires ABILHAND dans les 10 ordres aléatoires (10 feuilles)

**ABILHAND- Manual Ability Measure**

**French version**

Patient ______________________________________	Date ________________

|  | Quelle est la DIFFICULTE des activités suivantes ? | Impossible (0) | Difficile (1) | Facile (2) | ? |
| --- | --- | --- | --- | --- | --- |
| 1. | Soulever une casserole remplie |  |  |  |  |
| 2. | Faire le repassage |  |  |  |  |
| 3. | Ouvrir un paquet de chips |  |  |  |  |
| 4. | Tartiner une tranche de pain avec du beurre |  |  |  |  |
| 5. | Faire des pompes |  |  |  |  |
| 6. | Enfoncer un clou avec un marte |  |  |  |  |
| 7. | Ouvrir une boîte de conserve |  |  |  |  |
| 8. | Utiliser un tournevis |  |  |  |  |
| 9. | Décapsuler une bouteille |  |  |  |  |
| 10. | Secouer les draps |  |  |  |  |
| 11. | Ouvrir un bocal |  |  |  |  |
| 12. | Peler des pommes de terre avec un couteau |  |  |  |  |
| 13. | Tordre un torchon |  |  |  |  |
| 14. | Couper la haie |  |  |  |  |
| 15. | Faire les lacets |  |  |  |  |
| 16. | Pratiquer un sport de raquette |  |  |  |  |
| 17. | Applaudir vigoureusement |  |  |  |  |
| 18. | Manipuler le volant d'une voit |  |  |  |  |
| 19. | Se couper les ongles |  |  |  |  |
| 20. | Fermer la tirette d'une veste |  |  |  |  |
| 21. | Mettre des gants |  |  |  |  |
| 22. | Mélanger et distribuer des car |  |  |  |  |
| 23. | Essuyer les vitres |  |  |  |  |

Université catholique de Louvain & Haute Ecole Louvain en Hainaut ; [www.rehab-scales.org](http://www.rehab-scales.org)	Order 1

**ABILHAND- Manual Ability Measure**

**French version**

Patient ______________________________________	Date ________________

|  | Quelle est la DIFFICULTE des activités suivantes ? | Impossible (0) | Difficile (1) | Facile (2) | ? |
| --- | --- | --- | --- | --- | --- |
| 1. | Ouvrir une boîte de conserve |  |  |  |  |
| 2. | Fermer la tirette d'une veste |  |  |  |  |
| 3. | Se couper les ongles |  |  |  |  |
| 4. | Essuyer les vitres |  |  |  |  |
| 5. | Manipuler le volant d'une voit |  |  |  |  |
| 6. | Peler des pommes de terre avec un couteau |  |  |  |  |
| 7. | Utiliser un tournevis |  |  |  |  |
| 8. | Pratiquer un sport de raquette |  |  |  |  |
| 9. | Ouvrir un bocal |  |  |  |  |
| 10. | Mélanger et distribuer des car |  |  |  |  |
| 11. | Faire des pompes |  |  |  |  |
| 12. | Faire le repassage |  |  |  |  |
| 13. | Secouer les draps |  |  |  |  |
| 14. | Décapsuler une bouteille |  |  |  |  |
| 15. | Soulever une casserole remplie |  |  |  |  |
| 16. | Enfoncer un clou avec un marte |  |  |  |  |
| 17. | Couper la haie |  |  |  |  |
| 18. | Mettre des gants |  |  |  |  |
| 19. | Applaudir vigoureusement |  |  |  |  |
| 20. | Faire les lacets |  |  |  |  |
| 21. | Tartiner une tranche de pain avec du beurre |  |  |  |  |
| 22. | Ouvrir un paquet de chips |  |  |  |  |
| 23. | Tordre un torchon |  |  |  |  |

Université catholique de Louvain & Haute Ecole Louvain en Hainaut ; [www.rehab-scales.org](http://www.rehab-scales.org)	Order 2

**ABILHAND- Manual Ability Measure**

**French version**

Patient ______________________________________	Date ________________

|  | Quelle est la DIFFICULTE des activités suivantes ? | Impossible (0) | Difficile (1) | Facile (2) | ? |
| --- | --- | --- | --- | --- | --- |
| 1. | Peler des pommes de terre avec un couteau |  |  |  |  |
| 2. | Tordre un torchon |  |  |  |  |
| 3. | Ouvrir un paquet de chips |  |  |  |  |
| 4. | Décapsuler une bouteille |  |  |  |  |
| 5. | Utiliser un tournevis |  |  |  |  |
| 6. | Ouvrir un bocal |  |  |  |  |
| 7. | Se couper les ongles |  |  |  |  |
| 8. | Couper la haie |  |  |  |  |
| 9. | Manipuler le volant d'une voit |  |  |  |  |
| 10. | Faire le repassage |  |  |  |  |
| 11. | Ouvrir une boîte de conserve |  |  |  |  |
| 12. | Soulever une casserole remplie |  |  |  |  |
| 13. | Applaudir vigoureusement |  |  |  |  |
| 14. | Pratiquer un sport de raquette |  |  |  |  |
| 15. | Essuyer les vitres |  |  |  |  |
| 16. | Enfoncer un clou avec un marte |  |  |  |  |
| 17. | Mettre des gants |  |  |  |  |
| 18. | Tartiner une tranche de pain avec du beurre |  |  |  |  |
| 19. | Mélanger et distribuer des car |  |  |  |  |
| 20. | Faire des pompes |  |  |  |  |
| 21. | Secouer les draps |  |  |  |  |
| 22. | Faire les lacets |  |  |  |  |
| 23. | Fermer la tirette d'une veste |  |  |  |  |

Université catholique de Louvain & Haute Ecole Louvain en Hainaut ; [www.rehab-scales.org](http://www.rehab-scales.org)	Order 3

**ABILHAND- Manual Ability Measure**

**French version**

Patient ______________________________________	Date ________________

|  | Quelle est la DIFFICULTE des activités suivantes ? | Impossible (0) | Difficile (1) | Facile (2) | ? |
| --- | --- | --- | --- | --- | --- |
| 1. | Enfoncer un clou avec un marte |  |  |  |  |
| 2. | Utiliser un tournevis |  |  |  |  |
| 3. | Fermer la tirette d'une veste |  |  |  |  |
| 4. | Ouvrir une boîte de conserve |  |  |  |  |
| 5. | Faire des pompes |  |  |  |  |
| 6. | Pratiquer un sport de raquette |  |  |  |  |
| 7. | Mélanger et distribuer des car |  |  |  |  |
| 8. | Couper la haie |  |  |  |  |
| 9. | Mettre des gants |  |  |  |  |
| 10. | Se couper les ongles |  |  |  |  |
| 11. | Tartiner une tranche de pain avec du beurre |  |  |  |  |
| 12. | Faire les lacets |  |  |  |  |
| 13. | Tordre un torchon |  |  |  |  |
| 14. | Ouvrir un bocal |  |  |  |  |
| 15. | Ouvrir un paquet de chips |  |  |  |  |
| 16. | Manipuler le volant d'une voit |  |  |  |  |
| 17. | Peler des pommes de terre avec un couteau |  |  |  |  |
| 18. | Décapsuler une bouteille |  |  |  |  |
| 19. | Secouer les draps |  |  |  |  |
| 20. | Applaudir vigoureusement |  |  |  |  |
| 21. | Soulever une casserole remplie |  |  |  |  |
| 22. | Faire le repassage |  |  |  |  |
| 23. | Essuyer les vitres |  |  |  |  |

Université catholique de Louvain & Haute Ecole Louvain en Hainaut ; [www.rehab-scales.org](http://www.rehab-scales.org)	Order 4

**ABILHAND- Manual Ability Measure**

**French version**

Patient ______________________________________	Date ________________

|  | Quelle est la DIFFICULTE des activités suivantes ? | Impossible (0) | Difficile (1) | Facile (2) | ? |
| --- | --- | --- | --- | --- | --- |
| 1. | Manipuler le volant d'une voit |  |  |  |  |
| 2. | Ouvrir un paquet de chips |  |  |  |  |
| 3. | Tartiner une tranche de pain avec du beurre |  |  |  |  |
| 4. | Faire les lacets |  |  |  |  |
| 5. | Ouvrir un bocal |  |  |  |  |
| 6. | Faire des pompes |  |  |  |  |
| 7. | Fermer la tirette d'une veste |  |  |  |  |
| 8. | Utiliser un tournevis |  |  |  |  |
| 9. | Couper la haie |  |  |  |  |
| 10. | Applaudir vigoureusement |  |  |  |  |
| 11. | Essuyer les vitres |  |  |  |  |
| 12. | Ouvrir une boîte de conserve |  |  |  |  |
| 13. | Tordre un torchon |  |  |  |  |
| 14. | Faire le repassage |  |  |  |  |
| 15. | Enfoncer un clou avec un marte |  |  |  |  |
| 16. | Peler des pommes de terre avec un couteau |  |  |  |  |
| 17. | Pratiquer un sport de raquette |  |  |  |  |
| 18. | Se couper les ongles |  |  |  |  |
| 19. | Soulever une casserole remplie |  |  |  |  |
| 20. | Mélanger et distribuer des car |  |  |  |  |
| 21. | Décapsuler une bouteille |  |  |  |  |
| 22. | Secouer les draps |  |  |  |  |
| 23. | Mettre des gants |  |  |  |  |

Université catholique de Louvain & Haute Ecole Louvain en Hainaut ; [www.rehab-scales.org](http://www.rehab-scales.org)	Order 5

**ABILHAND- Manual Ability Measure**

**French version**

Patient ______________________________________	Date ________________

|  | Quelle est la DIFFICULTE des activités suivantes ? | Impossible (0) | Difficile (1) | Facile (2) | ? |
| --- | --- | --- | --- | --- | --- |
| 1. | Couper la haie |  |  |  |  |
| 2. | Tordre un torchon |  |  |  |  |
| 3. | Ouvrir une boîte de conserve |  |  |  |  |
| 4. | Mélanger et distribuer des car |  |  |  |  |
| 5. | Soulever une casserole remplie |  |  |  |  |
| 6. | Ouvrir un paquet de chips |  |  |  |  |
| 7. | Fermer la tirette d'une veste |  |  |  |  |
| 8. | Peler des pommes de terre avec un couteau |  |  |  |  |
| 9. | Secouer les draps |  |  |  |  |
| 10. | Faire des pompes |  |  |  |  |
| 11. | Applaudir vigoureusement |  |  |  |  |
| 12. | Faire les lacets |  |  |  |  |
| 13. | Utiliser un tournevis |  |  |  |  |
| 14. | Tartiner une tranche de pain avec du beurre |  |  |  |  |
| 15. | Ouvrir un bocal |  |  |  |  |
| 16. | Mettre des gants |  |  |  |  |
| 17. | Manipuler le volant d'une voit |  |  |  |  |
| 18. | Pratiquer un sport de raquette |  |  |  |  |
| 19. | Enfoncer un clou avec un marte |  |  |  |  |
| 20. | Essuyer les vitres |  |  |  |  |
| 21. | Faire le repassage |  |  |  |  |
| 22. | Se couper les ongles |  |  |  |  |
| 23. | Décapsuler une bouteille |  |  |  |  |

Université catholique de Louvain & Haute Ecole Louvain en Hainaut ; [www.rehab-scales.org](http://www.rehab-scales.org)	Order 6

**ABILHAND- Manual Ability Measure**

**French version**

Patient ______________________________________	Date ________________

|  | Quelle est la DIFFICULTE des activités suivantes ? | Impossible (0) | Difficile (1) | Facile (2) | ? |
| --- | --- | --- | --- | --- | --- |
| 1. | Mélanger et distribuer des car |  |  |  |  |
| 2. | Essuyer les vitres |  |  |  |  |
| 3. | Enfoncer un clou avec un marte |  |  |  |  |
| 4. | Ouvrir un paquet de chips |  |  |  |  |
| 5. | Manipuler le volant d'une voit |  |  |  |  |
| 6. | Ouvrir une boîte de conserve |  |  |  |  |
| 7. | Mettre des gants |  |  |  |  |
| 8. | Secouer les draps |  |  |  |  |
| 9. | Applaudir vigoureusement |  |  |  |  |
| 10. | Se couper les ongles |  |  |  |  |
| 11. | Faire le repassage |  |  |  |  |
| 12. | Tordre un torchon |  |  |  |  |
| 13. | Ouvrir un bocal |  |  |  |  |
| 14. | Décapsuler une bouteille |  |  |  |  |
| 15. | Fermer la tirette d'une veste |  |  |  |  |
| 16. | Soulever une casserole remplie |  |  |  |  |
| 17. | Couper la haie |  |  |  |  |
| 18. | Utiliser un tournevis |  |  |  |  |
| 19. | Pratiquer un sport de raquette |  |  |  |  |
| 20. | Faire les lacets |  |  |  |  |
| 21. | Peler des pommes de terre avec un couteau |  |  |  |  |
| 22. | Faire des pompes |  |  |  |  |
| 23. | Tartiner une tranche de pain avec du beurre |  |  |  |  |

Université catholique de Louvain & Haute Ecole Louvain en Hainaut ; [www.rehab-scales.org](http://www.rehab-scales.org)	Order 7

**ABILHAND- Manual Ability Measure**

**French version**

Patient ______________________________________	Date ________________

|  | Quelle est la DIFFICULTE des activités suivantes ? | Impossible (0) | Difficile (1) | Facile (2) | ? |
| --- | --- | --- | --- | --- | --- |
| 1. | Utiliser un tournevis |  |  |  |  |
| 2. | Manipuler le volant d'une voit |  |  |  |  |
| 3. | Se couper les ongles |  |  |  |  |
| 4. | Décapsuler une bouteille |  |  |  |  |
| 5. | Tartiner une tranche de pain avec du beurre |  |  |  |  |
| 6. | Faire le repassage |  |  |  |  |
| 7. | Secouer les draps |  |  |  |  |
| 8. | Ouvrir un paquet de chips |  |  |  |  |
| 9. | Enfoncer un clou avec un marte |  |  |  |  |
| 10. | Soulever une casserole remplie |  |  |  |  |
| 11. | Ouvrir un bocal |  |  |  |  |
| 12. | Mélanger et distribuer des car |  |  |  |  |
| 13. | Tordre un torchon |  |  |  |  |
| 14. | Peler des pommes de terre avec un couteau |  |  |  |  |
| 15. | Fermer la tirette d'une veste |  |  |  |  |
| 16. | Mettre des gants |  |  |  |  |
| 17. | Applaudir vigoureusement |  |  |  |  |
| 18. | Essuyer les vitres |  |  |  |  |
| 19. | Faire les lacets |  |  |  |  |
| 20. | Couper la haie |  |  |  |  |
| 21. | Pratiquer un sport de raquette |  |  |  |  |
| 22. | Faire des pompes |  |  |  |  |
| 23. | Ouvrir une boîte de conserve |  |  |  |  |

Université catholique de Louvain & Haute Ecole Louvain en Hainaut ; [www.rehab-scales.org](http://www.rehab-scales.org)	Order 8

**ABILHAND- Manual Ability Measure**

**French version**

Patient ______________________________________	Date ________________

|  | Quelle est la DIFFICULTE des activités suivantes ? | Impossible (0) | Difficile (1) | Facile (2) | ? |
| --- | --- | --- | --- | --- | --- |
| 1. | Faire les lacets |  |  |  |  |
| 2. | Applaudir vigoureusement |  |  |  |  |
| 3. | Enfoncer un clou avec un marte |  |  |  |  |
| 4. | Couper la haie |  |  |  |  |
| 5. | Faire des pompes |  |  |  |  |
| 6. | Mettre des gants |  |  |  |  |
| 7. | Peler des pommes de terre avec un couteau |  |  |  |  |
| 8. | Soulever une casserole remplie |  |  |  |  |
| 9. | Tordre un torchon |  |  |  |  |
| 10. | Secouer les draps |  |  |  |  |
| 11. | Ouvrir un bocal |  |  |  |  |
| 12. | Ouvrir un paquet de chips |  |  |  |  |
| 13. | Tartiner une tranche de pain avec du beurre |  |  |  |  |
| 14. | Faire le repassage |  |  |  |  |
| 15. | Décapsuler une bouteille |  |  |  |  |
| 16. | Fermer la tirette d'une veste |  |  |  |  |
| 17. | Essuyer les vitres |  |  |  |  |
| 18. | Utiliser un tournevis |  |  |  |  |
| 19. | Manipuler le volant d'une voit |  |  |  |  |
| 20. | Pratiquer un sport de raquette |  |  |  |  |
| 21. | Ouvrir une boîte de conserve |  |  |  |  |
| 22. | Mélanger et distribuer des car |  |  |  |  |
| 23. | Se couper les ongles |  |  |  |  |

Université catholique de Louvain & Haute Ecole Louvain en Hainaut ; [www.rehab-scales.org](http://www.rehab-scales.org)	Order 9

**ABILHAND- Manual Ability Measure**

**French version**

Patient ______________________________________	Date ________________

|  | Quelle est la DIFFICULTE des activités suivantes ? | Impossible (0) | Difficile (1) | Facile (2) | ? |
| --- | --- | --- | --- | --- | --- |
| 1. | Enfoncer un clou avec un marte |  |  |  |  |
| 2. | Essuyer les vitres |  |  |  |  |
| 3. | Applaudir vigoureusement |  |  |  |  |
| 4. | Faire les lacets |  |  |  |  |
| 5. | Ouvrir une boîte de conserve |  |  |  |  |
| 6. | Faire des pompes |  |  |  |  |
| 7. | Utiliser un tournevis |  |  |  |  |
| 8. | Mélanger et distribuer des car |  |  |  |  |
| 9. | Fermer la tirette d'une veste |  |  |  |  |
| 10. | Mettre des gants |  |  |  |  |
| 11. | Secouer les draps |  |  |  |  |
| 12. | Pratiquer un sport de raquette |  |  |  |  |
| 13. | Ouvrir un bocal |  |  |  |  |
| 14. | Tartiner une tranche de pain avec du beurre |  |  |  |  |
| 15. | Couper la haie |  |  |  |  |
| 16. | Se couper les ongles |  |  |  |  |
| 17. | Soulever une casserole remplie |  |  |  |  |
| 18. | Ouvrir un paquet de chips |  |  |  |  |
| 19. | Peler des pommes de terre avec un couteau |  |  |  |  |
| 20. | Faire le repassage |  |  |  |  |
| 21. | Manipuler le volant d'une voit |  |  |  |  |
| 22. | Tordre un torchon |  |  |  |  |
| 23. | Décapsuler une bouteille |  |  |  |  |

Université catholique de Louvain & Haute Ecole Louvain en Hainaut ; [www.rehab-scales.org](http://www.rehab-scales.org)	Order 10
